# Alterations in *PTPN11* and other RAS-/MAP-Kinase pathway genes define ganglioglioma with adverse clinical outcome and atypic histopathological features

**DOI:** 10.1101/2022.11.20.22282502

**Authors:** Lucas Hoffmann, Roland Coras, Katja Kobow, Javier A. López-Rivera, Dennis Lal, Costin Leu, Imad Najm, Peter Nürnberg, Jochen Herms, Patrick N. Harter, Christian G. Bien, Thilo Kalbhenn, Markus Müller, Tom Pieper, Till Hartlieb, Manfred Kudernatsch, Hajo Hamer, Sebastian Brandner, Karl Rössler, Ingmar Blümcke, Samir Jabari

## Abstract

The *PTPN11* gene was recently described as a novel lesional epilepsy gene by extensive exome-wide sequencing studies. However, germline mutations of *PTPN11* and other *RAS-/MAP*-Kinase signaling pathway *genes* cause Noonan syndrome, a multisystem disorder characterized by abnormal facial features, developmental delay, and sporadically, also brain tumors. Herein, we performed a deep phenotype-genotype analysis of a comprehensive series of ganglioglioma (GG) with brain somatic alterations of the *PTPN11* gene compared to GG with other common MAP-Kinase signaling pathway alterations. Seventy-two GG were submitted to whole exome sequencing and genotyping and 86 low grade epilepsy associated tumors (LEAT) to DNA-methylation analysis. Clinical data were retrieved from hospital files including postsurgical disease onset, age at surgery, brain localization, and seizure outcome. A comprehensive histopathology staining panel was available in all cases. We identified eight GG with *PTPN11* alterations, copy number variant (CNV) gains of chromosome 12, and the commonality of additional CNV gains in *FGFR4, RHEB, NF1, KRAS* as well as *BRAFV600E* alterations. Histopathology revealed an atypical and complex glio-neuronal phenotype with subpial tumor spread and large, pleomorphic, and multinuclear cellular features. Only three out of eight patients with GG and *PTPN11* alterations were free of disabling-seizures two years after surgery (38% Engel I). This was remarkably different from our series of GG with *BRAFV600E* mutations (85% Engel I). Our data point to a subgroup of GG with cellular atypia in glial and neuronal cell components, adverse postsurgical outcome, and genetically characterized by *PTPN11* and other Noonan syndrome-related alterations of the *RAS-/MAP*-Kinase signaling pathway. These findings need prospective validation in clinical practice as they argue for an adapted WHO grading system in developmental, glio-neuronal tumors associated with early-onset focal epilepsy. These findings also open avenues for targeted medical treatment.

## Introduction

Protein Tyrosine Phosphatase Non-receptor Type 11 (*PTPN11*) has been recently discovered as a new candidate gene in brain tissue obtained from drug-resistant structural epilepsies [2, 21] The *PTPN11* gene encodes for an early non-receptor-type protein tyrosine phosphatase SHP-2 (src homology region 2-domain phosphatase-2) of the RAS-/MAP-Kinase pathway. Germline variants in *PTPN11*, i.e., missense variants or copy number variations (CNV), or other *RAS-/MAP*-Kinase signaling pathway genes including *SHOC2, CBL, KRAS*, are known to cause an autosomal dominant multisystem disorder – the Noonan syndrome – or Noonan syndrome-associated disorders [10, 25, 28, 29]. Patients with Noonan syndrome sporadically develop glial and glio-neuronal brain tumors, like pilocytic astrocytoma and dysembryoplastic neuroepithelial tumors (DNT) [25, 28], together with several non-central nervous system (CNS) disorders, including cardiovascular abnormalities, lymphatic abnormalities, and growth hormone deficiencies.

We recently identified an accumulation of *PTPN11* alterations in low-grade epilepsy-associated brain tumors (LEAT) [21], the second largest lesion category in drug-resistant focal epilepsies amenable to neurosurgical treatment [6, 19]. Ganglioglioma (GG) account for approx. 64% of all LEAT, 82% of which affect the temporal lobe, and can mainly be histopathologically classified as WHO CNS grade 1 [30]. 80% of these patients become free from disabling seizures, i.e., five years after surgery, with many patients also tapering the anti-seizure medication [19]. Nevertheless, no biomarker is available for the remaining 20% of patients who do not benefit from a tailored epilepsy surgery approach.

We addressed this issue by studying a comprehensive cohort of GG with whole-exome sequencing, DNA methylation, histopathology, and clinical outcome parameter and identified gains in copy number variation and gain of function mutation of the *PTPN11* gene locus as a valuable predictor for adverse surgical outcome in patients with drug-resistant focal epilepsy and GG.

## Methods

One-hundred-thirty samples of confirmed GG and a pre-defined set of clinical features were collected at the University Hospital in Erlangen, Germany; Klinikum Bethel-Mara, Bielefeld University, Germany; and Schoen-Klinik Vogtareuth, Germany. Seventy-two samples were snap frozen at -80°C and submitted to whole-exome sequencing and Single-Nucleotide-Polymorphism analysis (SNP) with Global Screening Array Microarrays v1.0 (Illumina, San Diego, CA, USA) as described elsewhere [21]. Eighty-six samples were formalin-fixed and paraffin-embedded (FFPE) and submitted to DNA methylation analysis using 450K and 850K/EPIC arrays (Illumina, California, USA). Whole exome sequencing, SNP array, and DNA methylation analysis were jointly available in 28 samples (Table 1). The Ethics Committee of the Medical Faculty of the Friedrich-Alexander-University (FAU) Erlangen-Nürnberg, Germany, approved this study within the framework of the EU project “DESIRE” (FP7, grant agreement #602,531; AZ 92_14B) and European Reference Network EpiCARE” (grant agreement #769,051; AZ 193_18B).

### Histology and Immunohistochemistry

FFPE tissue blocks and glass slides were retrieved from the archives of the Neuropathological Institute at University Hospital Erlangen. Hematoxylin and Eosin stainings were available from all blocks and microscopically examined by two experienced neuropathologists (IB, RC). Additional immunohistochemical stainings were performed with the Ventana BenchMark ULTRA Immunostainer and the OptiView Universal DAB Detection Kit (Ventana Medical Systems, Tucson, AZ, USA) using the following panel of antibodies: Microtubule Associated Protein 2 (MAP2, clone C, mouse monoclonal, 1:100 dilution, Riederer Lausanne, Waadt, Swiss), Neuronal Nuclei (NeuN, clone A60, mouse monoclonal, 1:1500 dilution, Merck Millipore, Burlington, MA, USA), Kiel 67 protein (Mib1/Ki67, clone SP6/Ki67, rabbit monoclonal, 1:200 dilution, Cell Marque, Rocklin, CA, USA), glial fibrillary acid protein (GFAP, clone 6F2, mouse monoclonal, 1:500 dilution, Dako, Santa Clara, CA, USA), Synaptophysin (clone SP11, 1:100 dilution, Thermo Scientific, Waltham, MA, USA), isocitrate dehydrogenase-1 (IDH1, clone H09, mouse monoclonal, 1:50 dilution, Dianova, Eching, Bavaria, Germany), tumor protein p53 (p53, clone D0-7, mouse monoclonal, 1:2000 dilution, Dako, Santa Clara, CA, USA), Cyclin-dependent kinase inhibitor 2A (p16, clone G175-405, mouse monoclonal IgG1) and cluster of differentiation 34 (CD34, clone QBEnd-10, mouse monoclonal, 1:100 dilution, Dako, Santa Clara, CA, USA). Both reviewers unanimously achieved a histopathology diagnosis using the WHO classification of tumors of the CNS from 2016 and 2021.

### Whole-exome sequencing (WES) and copy-number variation (CNV) analysis

RC and IB histopathologically confirmed the presence of lesional cells in all frozen tissue samples (n=72). Following routine DNA extraction from the frozen tissue samples, whole-exome sequencing (WES) was performed at a coverage of >350x using Agilent SureSelect Human All Exon V7 enrichment and paired-end reads (151bp) Illumina sequencing. Paired-end FASTQ files were pre-processed following GATK best practices [13] and genetic variants were identified as described previously [21]. These samples were also genotyped using the Global Screening Array with Multi-disease drop-in (GSA-MD v1.0; Illumina, San Diego, CA, USA). The resulting single nucleotide polymorphism data was used to detect somaticsomatic CNV as described previously [21]. Here, genetic variants were defined as either mutation, CNV or single nucleotide polymorphism (SNP). The identified genetic variants were visualized with semi-automatically generated oncoplot graphics using Excel sheets 2016 oncoplot template (GitHub: https://github.com/ptgrogan/excel-oncoplot). The oncoplot was then sorted by genetic variants in the RAS-/MAP-Kinase signaling pathway to compare with histopathology features and clinical outcome data.

### DNA methylation array processing and CNV-calling

DNA was extracted from 86 FFPE tissue blocks histopathologically reviewed by RC and IB to confirm the presence of lesional cells using the QIAamp DNA Micro Kit (Qiagen, Venlo, Netherlands) following the manufacturer’s routine protocol. Methylation profiling was performed with the Infinium HumanMethylation450K in 19 samples (Table 1) or Infinium MethylationEPIC (EPIC) BeadChip in 67 samples (Illumina, San Diego, CA, USA). Twenty-eight cases had both, WES/SNP and DNA methylation analyses. In this study, we also included previously published methylation array data of 168 cases publically available from Capper et al. 2018 [8], and 26 cases by Wefers et al. 2020 [36].

We performed differential DNA methylation analysis using a self-customized Python wrapped cross R package pipeline as previously described [17] and publicly available at https://github.com/FAU-DLM/Methylr2py. Methylation data from 850K/EPIC and 450K data were combined into a virtual array with the ‘combineArrays’ function of the minfi package [1]. We stratified quantile normalized data using the ‘minfi’ ‘preprocessQuantile’ function [15]. Probes targeting sex chromosomes, containing single-nucleotide polymorphisms, not uniquely matching, and known cross-reactive probes were removed [11]. As a result, 128,525 probes were included in the combined virtual array and used for further analysis. Uniform Manifold Approximation and Projection (UMAP) for general non-linear dimensionality reduction was used for visualization [24] and to find subgroups compared to previously published LEAT cases [8]. The following non-default parameters were used: init = random, min_dist = 0.0, spread = 3.0.

To confirm the identified clusters from the previous step, we applied unsupervised learning using HDBSCAN as a clustering algorithm [23]. The following non-default parameters were used herein: min_samples = 4, min_cluster_size = 4.

Subsequent copy number calling was performed with ‘conumee’ Bioconductor package v.1.28.0. [16]. To be able to perform summary plots of identified clusters from the HDBSCAN, we adopted and extended the ‘conumee’ package functionality. The new functions enabling summary plots from the ‘conumee’ package are forked from the original project and publicly available at https://github.com/FAU-DLM/conumee.

## Results

### Whole exome sequencing and CNV detection

Out of 72 samples of GG submitted to WES, 44 carried a genetic variant (61%), i.e., mutations, gains in copy number variants (CNV), or loss of heterozygosity (LOH) and were further studied herein (**Fig. 1**). Our histopathology review confirmed the diagnosis of ganglioglioma CNS WHO grade 1 regarding histological and immunohistological criteria of the WHO classification of tumors of the CNS from 2016 and 2021. One case was compatible with the diagnosis of a multinodular and vacuolating tumor (MNVT; case #14). Quality assessment and significance levels of the genomic data were determined as described previously [21].

**Figure 1:**
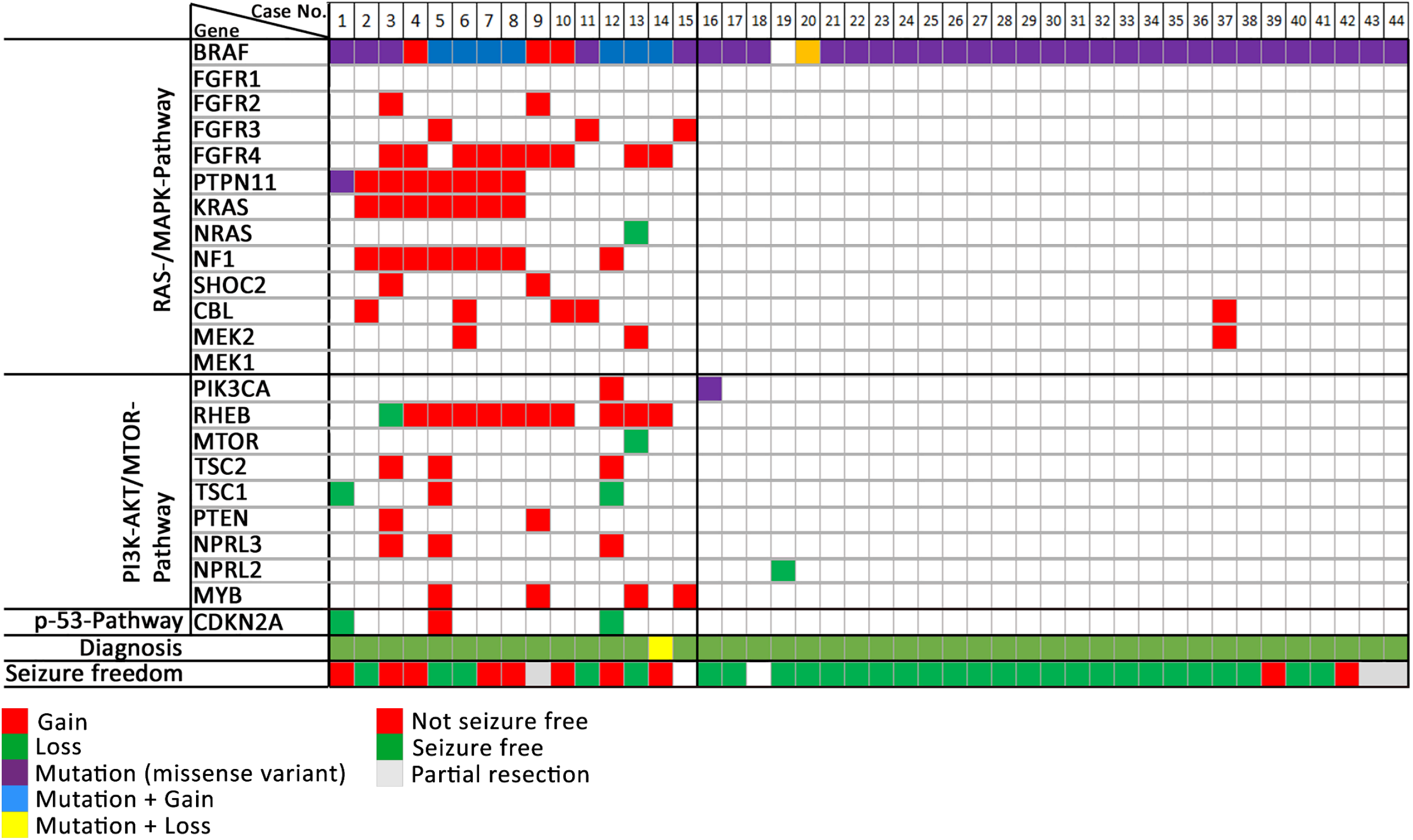
Oncoplot of WES and CNV detection. Each column represents a patient sample submitted to WES (same ID as in supplemental **Table 1**). We have intentionally classified these data into three groups of patients: GG with complex genetic variants (cases 1-15), GG with non-complex genetic variants (cases 16-44), and GG without variants (cases 45-72, not shown). **Diagnosis**: Histopathology review confirmed the diagnosis of ganglioglioma (green tiles) in 43 cases. Case #14 was compatible with the diagnosis of MNVT (yellow tile). **Seizure freedom** was defined by Engel’s outcome class 1 [19]. Lack of seizure freedom was explicable by incomplete resection in cases #9, #42, and #43 (greyish tiles). Outcome data was missing in cases #15 and #18 (white tiles).

We observed three major patterns of genetic abnormalities in this cohort. Group 1 consisted of 15 cases with complex genetic variants (CGV) and several parallel aberrations in the *RAS/MAP*-Kinase and *PI3K/AKT/MTOR* pathways. Eight samples had alterations in the *PTPN11* gene (53%), seven with CNV gains and one with a missense variant. These eight samples frequently had also mutations in *BRAFV600E* (n=7, 86%) and CNV gains in *KRAS* (n=7), *NF1* (n=7), *FGFR2*-4 (n=6). Less frequent CNV gains were observed in *BRAF* (n=5), *RHEB* (n=5), and other *RAS-/MAPK*-pathway genes (n=4). The remaining seven samples of this group of GG with complex and multiple hits all had a *BRAFV600E* mutation and/or *BRAF* gains plus additional hits in *FGFR2*-*4* (n=6, 86%), RHEB (n=5, 71%), *CBL* (n=2, 29%), or *SHOC2, MEK2, NF1*, (n=1 for each), *CDKN2A* LOH (n=1) and other *PI3K/AKT/MTOR* pathway genes (n=6, 86%).

Group 2 contained 29 tissue samples characterized by a *BRAFV600E* mutation (n=28; 97%) or other non-complex genetic alterations. Three samples had a *BRAFV600E* mutation and a CNV loss in *BRAF*, a *PIK3CA* missense variant, or a gain in *CBL* and *MEK2*. One sample only had LOH of *NPRL2* (**Fig. 1**).

The remaining 28 cases of our cohort of GG submitted to WES revealed no genetic alterations discernable by our techniques (group 3; 39%).

### DNA Methylation analysis

DNA methylation array data were obtained from 86 patients in our series (**Table 1**). In addition, we retrieved 194 reference samples from published sources to apply UMAP analysis and unsupervised clustering using the HDBSCAN methods. Additional copy-number profiling was performed utilizing the ‘conumee’ package of all the samples mentioned above.

UMAP and HDBSCAN analysis depicted readily expected and established methylation classes for the reference cohort. Twenty-one of our samples were also assigned to these established groups (**Fig. 2**; supplemental **Table 1**). This reference cohort consisted of Ganglioglioma (LGG, GG), Dysembryoplastic Neuroepithelial Tumours (LGG, DNT), Pleomorphic Xantoastrocytoma (LGG, PXA), and diffuse astrocytoma, MYB-or MYB-L1 altered (LGG, MYB). However, we detected a fifth, new methylation class, including all samples of our series with complex genetic variants (as shown in **Fig.1**), including those altered in *PTPN11*. Sixty-five samples were obtained from patients not being seizure-free after epilepsy surgery, i.e., adverse outcome, and samples with atypical histopathology also fell into this new DNA methylation class. This class also contained samples missing further molecular subtyping or clinical outcome data. Of these, most were histopathologically diagnosed as ganglioglioma (n=56), ganglioglioma with increased proliferation rate analog WHO II° (n=1), anaplastic ganglioglioma WHO III° (n=1), isomorphic astrocytoma (n=2), PXA (n=5).

**Figure 2:**
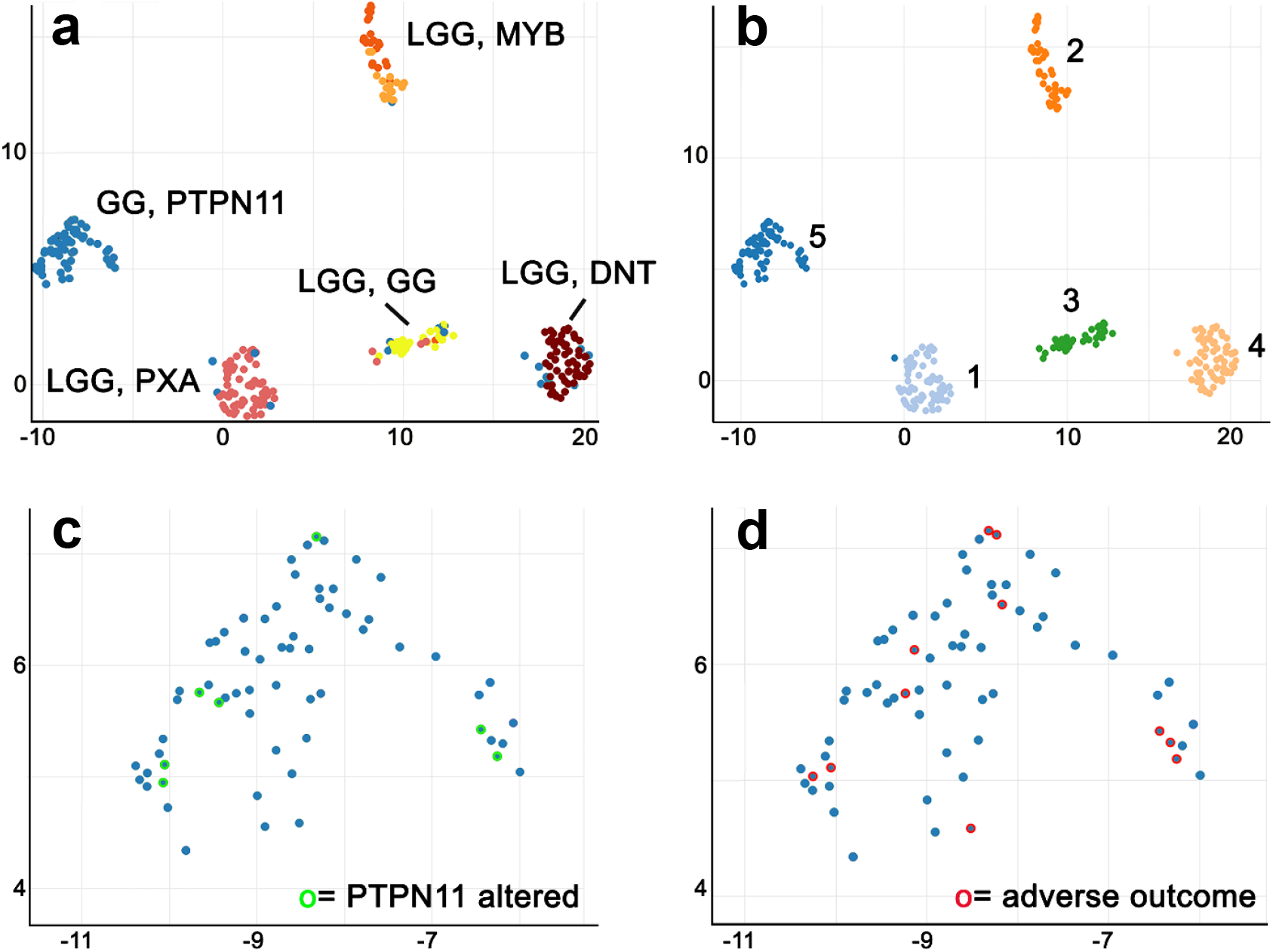
Methylome and CNV clustering defined LEAT subgroups. UMAP-clustering (**a**) of methylation data revealed a novel methylation class distinct from previously recognized groups (GG *PTPN11*; n=65). The reference groups were annotated as low-grade glioma, Ganglioglioma (LGG, GG, n=26 in yellow), Dysembryoplastic Neuroepithelial Tumour (LGG, DNT, n=56 in brown), Pleomorphic Xantoastrocytoma (LGG, PXA, n=63 in pinkish) and diffuse astrocytoma MYB-or MYB-L1 altered (LGG, MYB, n=45 in orange) by Capper et al. 2018 and Wefers et al. 2020. The separate Erlangen cohort is labeled in blue.**b:** Unsupervised clustering using HDBSCAN confirmed the novel DNA methylation class (class 5 on the left). Seven samples with *PTPN11* alterations were assigned to this group (green open circles in **c**). Eleven out of 19 patients not seizure-free in our cohort were also assigned to this group (red open circle in **d;** patients with partial resection and cases that have failed quality control were not visualized).

Unsupervised clustering by HDBSCAN confirmed the presence of a fifth methylation class distinct from previously recognized histopathology diagnosis, i.e., GG, DNT, PXA, and LGG, MYB altered.

### Copy number profiling

We performed an additional analysis of copy number calling from 450k and EPIC array data using the ‘conumee’ package to confirm the SNP-based CNV detection. Typical GG of the DNA methylation class LGG, GG showed a flat CNV profile with marginal gains at chromosomes 5, 7 and 12 (**Fig. 3**), compatible with *RAS-/MAP*-kinase pathway alterations described in this diagnostic entity [7]. In contrast, the new class comprising GG with complex genomic alterations and an adverse postsurgical outcome, i.e. GG *PTPN11*, showed frequent gains at chromosomes 5, 7, and 12, as well as losses at chromosomes 6, 7, 8, and partially of chromosome 9 (**Fig. 3 c, d**). The *PTPN11* altered samples accounted for most gains of chromosomes 5 and 12, i.e., *PTPN11* and other *RAS-/MAP*-kinase pathway genes located on chromosome 12, and the alterations of chromosome 7.

**Figure 3:**
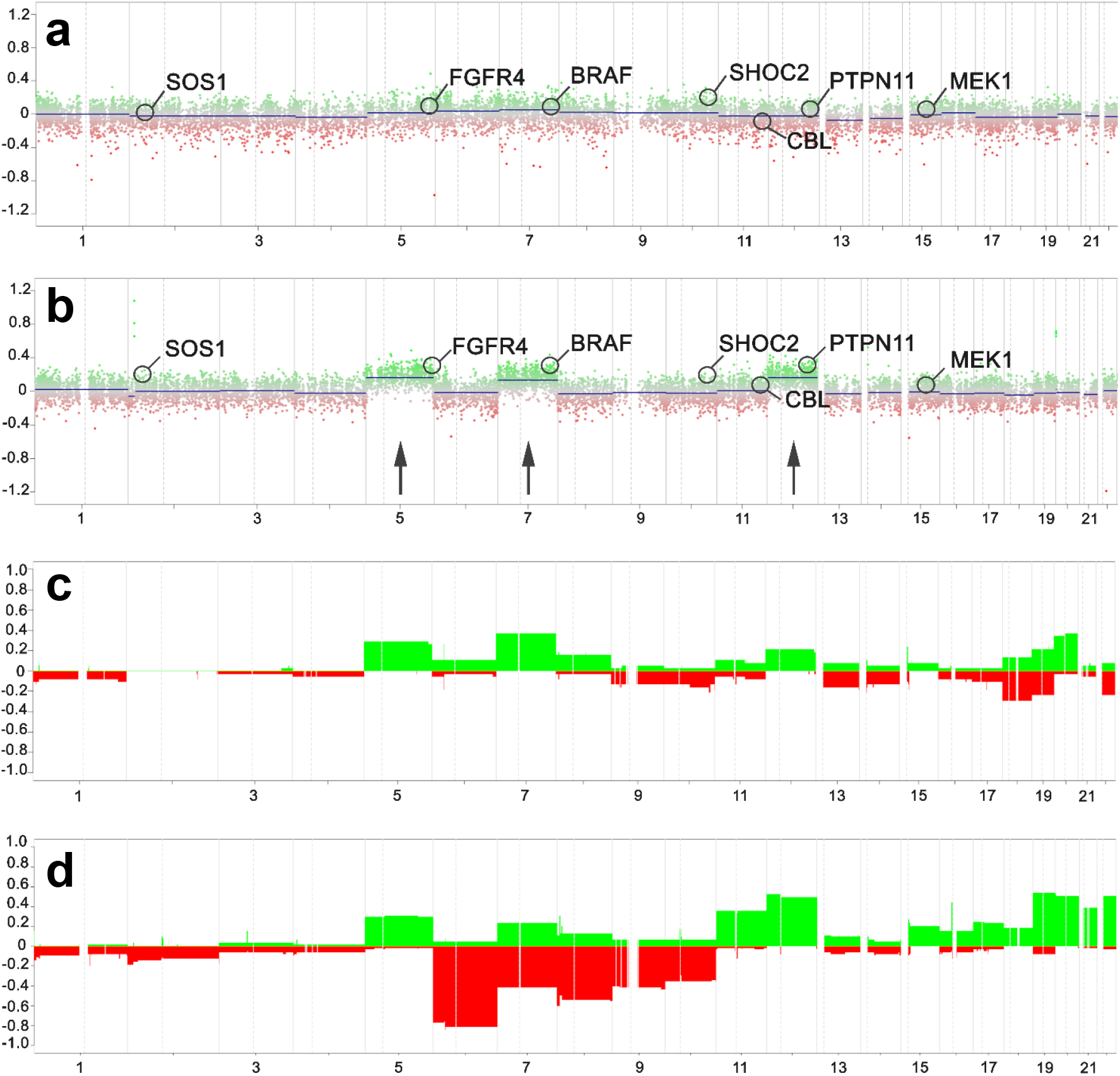
Copy number profile of different methylation groups. **CNV plots of representative samples and summary plots of ganglioglioma of the reference cohort and the adverse ganglioglioma methylation group** While the published reference cohort of ganglioglioma showed a flat profile with marginal gains at chromosomes 5, 7 and 12 in **a and c** (summary plot of entire cohort, n=38), frequent gains and losses were evident within the new ganglioglioma methylation class with adverse outcome in **b** (single patient sample #7, see supplemental Table 1) and **d** (summary plot of entire cohort, n=65). In summary plots, the y-axis indicated the relative share of alterations within the sample cohort.

### Genotype-phenotype correlations

We further assessed the histopathology phenotype of eight GG with *PTPN11* alterations of our cohort as identified and confirmed by molecular-genetic studies. These tumors were characterized by a glioneuronal phenotype (Figure 4). The neoplastically transformed glial cell population shared features of astroglia including prominent immunoreactivity with antibodies against GFAP (**Fig. 4 f**). Oligodendroglia-like cell features were not dominant in these tumors. Dysplastic neurons were another hallmark, some of which were located in the subarachnoidal space (**Fig. 4 e**) containing multiple nuclei and immunoreactivity for MAP2 and synaptophysin. Tumor growth was mostly diffuse into the grey and white matter (**Fig. 4 b**), but partially also nodular. Interestingly, seven tumors revealed growth into the subarachnoidal space (**Fig. 4 b**). Immunohistochemistry was most helpful recognizing the complex growth pattern of these tumors, which were immunoreactive for CD34 and p16 (**Fig. 4 c and d**). In contrast to p16, CD34 was consistently seen also in the subarachnoidal space. There was no evidence for mutant *IDH1 R132H*-specific labeling nor increased proliferation activity above 5%. Some specimens also showed small calcifications.

**Figure 4:**
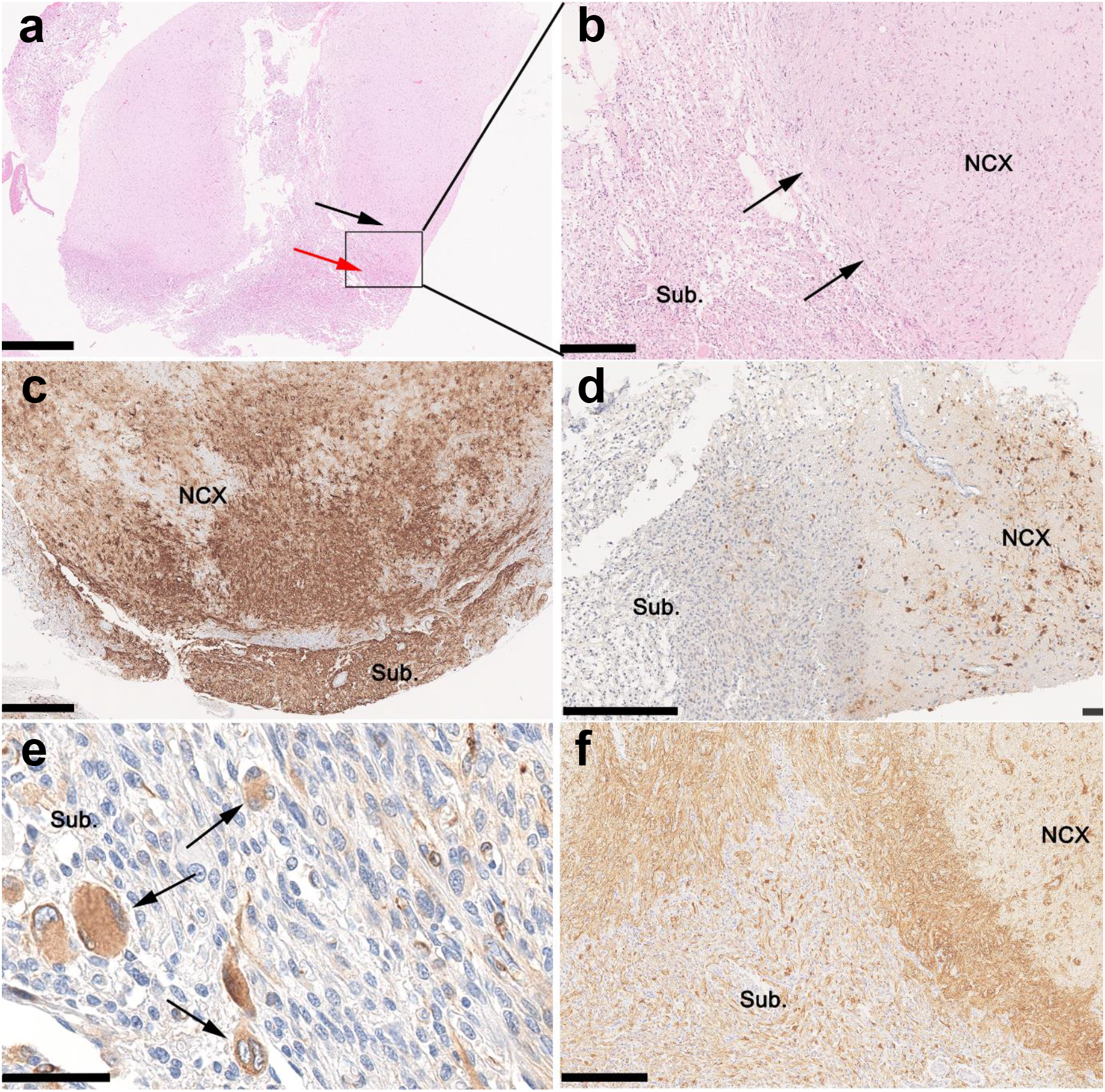
Histopathology findings in *PTPN11* altered Ganglioglioma. A representative case of *PTPN11* altered ganglioglioma (case 8; supplement Table 1). **a-b:** Intra- and extracortical tumor growth (black and red arrow in **a**, respectively) was readily visible on HE staining. Arrows in **b** indicate the border of the cortical surface and subarachnoidal space. **c:** CD34-immunoreactivity was a prominent feature of cortical and subarachnoidal tumor growth. **d:** In contrast, p16-immunoreactivity was accentuated in the cortical tumor component (subarachnoidal component on the right). **e:** Multinucleated cells in the subarachnoidal space immunoreactive for MAP2 were classified as dysplastic neurons (arrows) **f:** The astroglial component was predominant (GFAP immunohistochemistry). NCx – neocortex, SUB – subarachnoidal space. Scale bars: **a:** 1mm; **b, d, f**: 250μm; **c**: 500μm; **e:** 100μm.

*PTPN11*-altered tumors accounted for 53% of tumors with complex genetic variants. Interestingly, five out of eight patients with *PTPN11* altered GG were not seizure-free after surgery nor tapered off antiseizure medication. Furthermore, one patient died of sudden unexpected death in epilepsy (SUDEP; supplemental **Table 1**). The low seizure freedom rate of 38% Engel class I in this cohort contrasts with the excellent outcome in 76% of patients with GG characterized just by *BRAFV600E* mutations. This holds true also when comparing the group of all GG with complex genetic variants (n=15) with group 2 (n=19 had available clinical information; Chi-square p = 0.007; Yates correction p = 0.018). Sufficient clinical data within the group of GG without any *genetic varian*t was available in 22 out of 27 cases, of which 72% were seizure-free after surgery. 6 out of 22 (28,5%) were not completely seizure-free. One patient had a provoked seizure (alcohol + sleep deprivation; case 47) and another had a partial resection (case 46). However, all patients with sufficient follow-up data remained on antiseizure medication two years after surgery.

## Discussion

Our study identified a new DNA methylation class consisting of 65 cases of patients with LEAT submitted to epilepsy surgery due to intractable focal seizures. Seven of the eight *PTPN11-*altered GG recognized in this study were assigned to the novel DNA methylation class by the algorithm. 62% of these patients had adverse postsurgical outcomes, i.e., not seizure-free, 88% had subarachnoidal growth of biphasic, glio-neuronal cells, and all carried complex brain somatic gene variants including the *RAS-/MAP*-Kinase and PIK3-AKT/MTORpathways. These findings warrant further analysis in larger and prospectively collected tumor cohorts as patients will require adjunct medical therapies to prevent the harmful consequences of active, long-term epilepsy.

The histopathological classification of LEAT remains an ever-challenging issue [3, 5, 30]. This applies in particular to the variable phenotypes of GG due to more or less dominant i) neoplastically transformed astroglia, ii) neoplastically transformed oligodendroglial-like cells and iii) dysplastic neuronal cell population [5]. Subarachnoidal growth and adverse outcome in our tumor cohort suggested the differential diagnosis of PXA. However, neither the histopathology phenotype, e.g., the absence of reticulin fibers and xanthomatous cells, nor the absence of homozygous deletions of CDKN2A/B supported this diagnosis in our cohort (**Fig. 3 and 4**). In addition, DNA methylation profiling readily separated the class of *PTPN11*-altered GG from PXA (**Fig. 2**). Several publications describe the composite tumors of variable entities, most often including GG and PXA [22, 35], a feature in need of further and systematic exploration in the LEAT cohort.

As proven for many high-grade gliomas and embryonal brain tumours described in the 5^th^ edition of the WHO classification scheme, comprehensive genotype-phenotype studies are likely to resolve the issue of interrater reliability and agreement, when based on robust clinical data. We applied genomewide deep sequencing combined with CNV and DNA methylation array analysis and microscopy studies of 72 snap-frozen GG obtained from patients with careful clinical characterization and postsurgical follow-up to address this issue. Interestingly, our study design identified a cohort of 65 GG as a novel DNA methylation class containing samples not recognizable with data used to build the Heidelberg classifier (**Fig. 3**). *PTPN11-* altered GG accumulated in this class and herein this class was provisionally termed *PTPN11-*altered GG.

*PTPN11* was previously identified as a novel epilepsy gene [2, 21]. However, it was never described before in brain somatic disorders. Germline alterations of *PTPN11* are associated with Noonan (NS, Figure 5), the most frequent dysfunctional growth syndrome. Neuroepithelial tumors also occur in patients with NS as does focal epilepsy [20, 28, 29, 32]. Other members of the *RAS-/MAP*-kinase pathway affected within our series of *PTPN11*-altered GG were also known to contribute to Noonan-associated syndromes, e.g., *SHOC2, CBL* or *MEK1* [25, 28, 29]. In our cohort they either revealed a gain of function mutation or a gain of copy numbers at the level of brain mosaicism [21]. However, experimental evidence is scarce for most genetic alterations in GG and focused on *BRAFV600E. Inutero* electroporation of *BRAFV600E* in the developing mouse brain confirmed the histogenesis of CD34 immunoreactive GG with focal epileptogenesis [18]. This finding aligns with our second cohort of 24 patients with GG and BRAFV600E (Figure 1). However, our cohort of *PTPN11*-altered GG also showed alterations in the *PI3K-AKT/MTOR* pathway frequently involved in epileptogenic Focal Cortical Dysplasia [4]. The newly described experimental ganglioglioma mouse model of *BRAFV600E* with mTOR pathway alterations would thus be an intriguing option to further explore the nature of *PTPN11*-altered GG [9].

**Figure 5:**
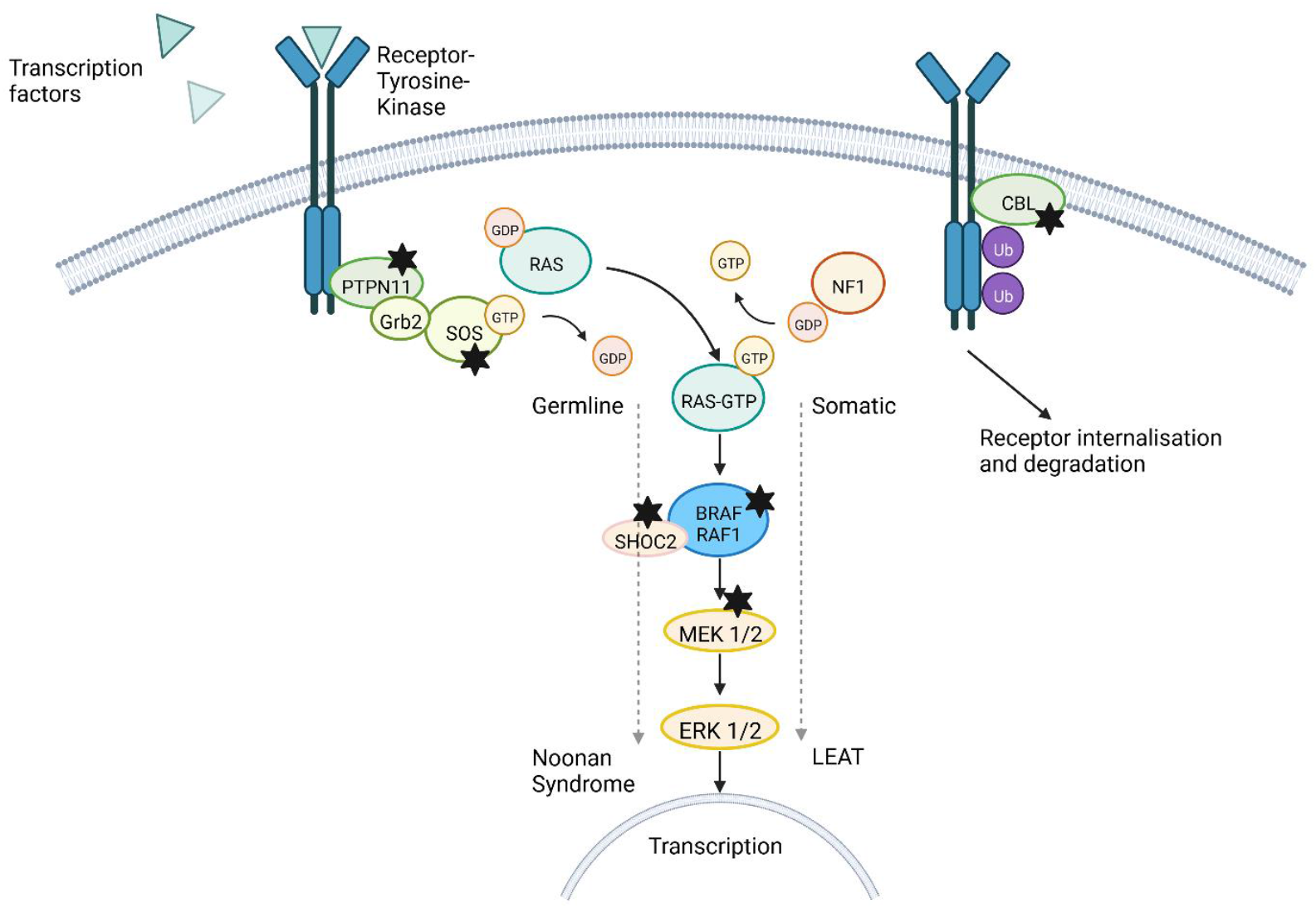
RAS-/MAP-Kinase pathway in GG, *PTPN11* altered. Germline alterations of *PTPN11* and other *RAS-/MAP*-kinase-pathway genes were known to cause Noonan-Syndrome [25, 28, 29]. Genes marked with a black star showed alterations in our cohort of GG.

It is difficult to reconcile the failure of seizure freedom following epilepsy surgery in our retrospective and small patient cohort. Subarachnoidal tumor growth might contribute to the incompleteness of surgical resection, particularly when not readily anticipated from pre-surgical neuroimaging studies. Another option is a larger tumor area not visible on MRI and not included in the surgical field as indicated by the diffuse infiltration pattern of CD34- and p16-immunoreactive tumor satellites. The impact of transcriptional signatures in LEAT reflecting the clinical outcome has also been recently postulated [12]. This work distinguished three subgroups of GG, i.e., *BRAF*-altered GG, juvenile GG, and not otherwise specified. Interestingly, the *BRAF*-altered group of GG was characterized by transcriptional changes in *RAS-/MAPK*-pathway including the *FGFR* genes. These cases also had a “progressive disease” after surgery including tumor relapse/recurrence and were postulated to aim for gross-total resection to prevent patients from tumor recurrence. However, the epileptogenic tumor network can also be compromised by other cell biological features, e.g., the microtubular glioma network, which is not yet studied nor anticipated well in LEAT [27, 33, 34].

Lack of seizure-freedom significantly impacts the quality of life and patient survival, i.e. sudden unexpected death in epilepsy (SUDEP); [14]. Many studies indicate that the overall mortality rate for people with epilepsy is elevated two- or threefold compared to the general population [26]. Indeed, our patient #8 (supplemental **Table 1, Fig. 1**) died from SUDEP two years after a seizure relapse from an otherwise successful surgical approach. Such adverse clinical features are not recognized in the WHO grading scale, which only addresses the risk of tumor recurrence and/or malignant progression. This contrasts routine clinical practice in the realm of epileptology and epilepsy surgery, in which seizure freedom is of utmost importance for patient survival and quality of life, in particular for the group of LEAT with their most successful outcome prediction measures [6, 19]. The advancement of comprehensive genotype-phenotype association studies may reinforce the discussion and communication across medical disciplines to help developing the best available disease classification schemes and reflecting and taking into account all relevant disease parameters.

As a matter of fact, integrated genotype-phenotype classification schemes increasingly govern targeted medical therapies. Fast access to tailored epilepsy surgery will remain, however, of utmost importance in patients with focal, drug-resistant seizures. Tumour tissue samples will then help to reliably identify a tumour subtype and possible molecular targets when surgery had failed. Indeed, a variety of targeted therapies were already developed for the gene product of the *PTPN11*, such as orthosteric inhibition of the *SHP2* protein [31].

## Data Availability

The complete methylation data of the 86 samples included in this study, will be deposited in NCBIs Gene Expression Omnibus (GEO http://www.ncbi.nlm.nih.gov/geo). The accession numbers follow.

## Data availability

The complete methylation data of the 86 samples included in this study, will be deposited in NCBI’s Gene Expression Omnibus (GEO, http://www.ncbi.nlm.nih.gov/geo). The accession number is GSE218542.

## Acknowledgments

The study was supported by the German Research Foundation DFG Bl421/4-1 to IB; NU 50/13-1 to PN, and LA 4193/2-1 to DL. This work was supported by the DFG Research Infrastructure West German Genome Center (project 407493903) as part of the Next Generation Sequencing Competence Network (project 423957469). NGS analyses were carried out at the production site Cologne (Cologne Center for Genomics). SJ is supported by the Interdisziplinäres Zentrum für Klinische Forschung, Universitätsklinkum Erlangen (IZKF; project number J81).

## Supplements

**Supplementary Table 1:** WES, SNP and methylation cohort. ID = case numbers, * = adverse outcome due to incomplete resection, ** = patient received a second surgery due to lack of seizure freedom; Platform: WES = Whole-Exome Sequencing, SNP = Single-Nucleotide-Polymorphism array, EPIC or 450k DNA methylation arrays; Outcome = most recent outcome according to Engel; N/A = not available; Follow-up in years; D. o. E. = Duration of Epilepsy; Onset= disease onset (age in years); Sex: F − female, M = male; DX = histopathology diagnosis; MC − methylation classes (see Figure 2).

| Case ID | Platform | Outcome | Follow-up (y) | D. o. E | Onset | Sex | DX | MC |
| --- | --- | --- | --- | --- | --- | --- | --- | --- |
| 1 | GSA-MA/WES + EPIC | Engel 4 | 4 | 2 | N/A | M | GG 1 | GG, PTPN11 |
| 2 | GSA-MA/WES + EPIC | Engel 1a | 2 | 2 | N/A | M | GG 1 | GG, PTPN11 |
| 3** | GSA-MA/WES + 450k | Engel 1b | 2 | 4 | 11-15 | M | GG 1 | LGG, PXA |
| 4 | GSA-MA/WES + EPIC | Engel 2b | 2 | 40 | 11-15 | F | GG 1 | GG, PTPN11 |
| 5 | GSA-MA/WES + EPIC | Engel 1a | 2 | 4 | 6-10 | M | GG 1 | GG, PTPN11 |
| 6 | GSA-MA/WES + EPIC | Engel 1a | 2 | 1 | 11-15 | F | GG 1 | GG, PTPN11 |
| 7 | GSA-MA/WES + EPIC | Engel 1c | 2 | 2 | 16-20 | F | GG 1 | GG, PTPN11 |
| 8 | GSA-MA/WES + EPIC | Engel 2d<br>SUDEP | 2 | 13 | 16-20 | F | GG 1 | GG, PTPN11 |
| 9 | GSA-MA/WES | Engel 1b* | 6 | 4 | 6-10 | F | GG 1 |  |
| 10 | GSA-MA/WES + EPIC | Engel 1b | 3 | 4 | 16-20 | F | GG 1 | GG, PTPN11 |
| 11 | GSA-MA/WES + EPIC | Engel 1a | 2 | 3 | 11-15 | M | GG 1 | GG, PTPN11 |
| 12** | GSA-MA/WES | Engel 1c | 2 | 5 | 16-20 | F | GG 1 |  |
| 13 | GSA-MA/WES + EPIC | Engel 1a | 2 | 0,3 | 6-10 | M | GG 1 | GG, PTPN11 |
| 14 | GSA-MA/WES | Engel 2b | 2 | 45 | 0-5 | M | MNVT |  |
| 15 | GSA-MA/WES | N/A | N/A | N/A | N/A | M | GG 1 |  |
| 16 | GSA-MA/WES | Engel 1a | 2 | 1,75 | 0-5 | M | GG 1 |  |
| 17 | GSA-MA/WES | Engel 1a | 2 | 8,75 | 0-5 | M | GG 1 |  |
| 18 | GSA-MA/WES | N/A | N/A | N/A | N/A | F | GG 1 |  |
| 19 | GSA-MA/WES + EPIC | Engel 1a | 2 | 13 | 6-10 | M | GG 1 | GG, PTPN11 |
| 20 | GSA-MA/WES | Engel 1a | 2 | 5 | 11-15 | M | PXA/<br>GG |  |
| 21 | GSA-MA/WES | Engel 1a | 2 | no seizures |  | F | GG 1 |  |
| 22 | GSA-MA/WES | Engel 1a | 2 | 5 | 0-5 | M | GG 1 |  |
| 23 | GSA-MA/WES | Engel 1a | 2 | 7 | 11-15 | M | GG 1 |  |
| 24 | GSA-MA/WES | Engel 1a | 2 | 3 | 0-5 | M | GG 1 |  |
| 25 | GSA-MA/WES | Engel 1a | 2 | 9 | 6-10 | F | GG 1 |  |
| 26 | GSA-MA/WES | Engel 1a | 2 | 1 | 6-10 | M | GG 1 |  |
| 27 | GSA-MA/WES | Engel 1a | 2 | 3 | 16-20 | M | GG 1 |  |
| 28 | GSA-MA/WES | Engel 1a | 2 | 7 | 6-10 | F | GG 1 |  |
| 29 | GSA-MA/WES | Engel 1a | 2 | 9 | 6-10 | F | GG 1 |  |
| 30 | GSA-MA/WES | Engel 1a | 2 | 1 | 0-5 | M | GG 1 |  |
| 31 | GSA-MA/WES +<br>EPIC | Engel 1a | 2 | N/A | N/A | M | GG 1 | GG, PTPN11 |
| 32 | GSA-MA/WES | Engel 1a | 2 | 3 | 0-5 | F | GG 1 |  |
| 33 | GSA-MA/WES | Engel 1a | 2 | 2,25 | 0-5 | F | GG 1 |  |
| 34 | GSA-MA/WES | Engel 1a | 2 | 6 | 11-15 | M | GG 1 |  |
| 35 | GSA-MA/WES | Engel 1a | 2 | 3 | 0-5 | M | GG 1 |  |
| 36 | GSA-MA/WES +<br>EPIC | Engel 1a | 2 | N/A | N/A | M | GG 1 | GG, PTPN11 |
| 37 | GSA-MA/WES +<br>EPIC | Engel 1a | 2 | 11 | 11-15 | F | GG 1 | GG, PTPN11 |
| 38 | GSA-MA/WES +<br>EPIC | Engel 1a | 2 | N/A | N/A | M | GG 1 | GG, PTPN11 |
| 39 | GSA-MA/WES +<br>EPIC | Engel 2a | 2 | 6 | 0-5 | M | GG 1 | GG, PTPN11 |
| 40 | GSA-MA/WES | Engel 1a | 2 | N/A | N/A | M | GG 1 |  |
| 41 | GSA-MA/WES +<br>EPIC | Engel 1a | 2 | 6 | 0-5 | M | GG 1 | GG, PTPN11 |
| 42 | GSA-MA/WES | Engel 3a | 2 | 2 | 0-5 | M | GG 1 |  |
| 43* | GSA-MA/WES +<br>EPIC | Engel 3a | 2 | 24 | 0-5 | F | GG 1 | GG, PTPN11 |
| 44 | GSA-MA/WES | Engel 1c | 2 | 6 | 0-5 | F | GG 1 |  |
| 45* | GSA-MA/WES | Engel 3a | 2 | 8 | 46-50 | F | GG 1 |  |
| 46* | GSA-MA/WES + EPIC | Engel 2b | 2 | 16 | 16-20 | M | GG 1 | GG, PTPN11 |
| 47** | GSA-MA/WES + EPIC | Engel 3a | 2 | 4 | 21-25 | F | GG 1 | GG, PTPN11 |
| 48 | GSA-MA/WES + EPIC | Engel 1b | 2 | 11 | 11-15 | M | GG 1 | GG, PTPN11 |
| 49 | GSA-MA/WES + EPIC | Engel 1b | 2 | 30 | 0-5 | F | GG 1 | GG, PTPN11 |
| 50 | GSA-MA/WES + EPIC | Engel 1b | 2 | 6 | 0-5 | M | GG 1 | GG, PTPN11 |
| 51 | GSA-MA/WES | Engel 1a | 2 | 4 | 6-10 | M | GG 1 |  |
| 52 | GSA-MA/WES | Engel 1a | 2 | 5 | 6-10 | F | GG 1 |  |
| 53 | GSA-MA/WES | Engel 1a | 2 | 14 | 11-15 | M | GG 1 |  |
| 54 | GSA-MA/WES | Engel 1a | 2 | 0 | 0-5 | M | GG 1 |  |
| 55 | GSA-MA/WES | Engel 1a | 2 | 13 | 26-30 | M | GG 1 |  |
| 56 | GSA-MA/WES + EPIC | Engel 1a | 2 | 12 | 26-30 | M | GG 1 | GG, PTPN11 |
| 57 | GSA-MA/WES | N/A | N/A | N/A | N/A | F | GG 1 |  |
| 58 | GSA-MA/WES + 450k | N/A | N/A | N/A | N/A | M | GG 1 | LGG, GG |
| 59 | GSA-MA/WES | N/A | N/A | N/A | N/A | M | GG 1 |  |
| 60 | GSA-MA/WES | Engel 1a | 2 | 24 | 0-5 | F | GG 1 |  |
| 61 | GSA-MA/WES | Engel 1a | 2 | 2 | 6-10 | M | GG 1 |  |
| 62 | GSA-MA/WES | Engel 1a | 2 | 0,25 | 0-5 | F | GG 1 |  |
| 63 | GSA-MA/WES | Engel 1a | 2 | 2 | 0-5 | F | GG 1 |  |
| 64 | GSA-MA/WES | Engel 1a | 2 | 4 | 0-5 | M | GG 1 |  |
| 65 | GSA-MA/WES | N/A | N/A | N/A | N/A | F | GG 1 |  |
| 66 | GSA-MA/WES | Engel 1a | 2 | 2 | 11-15 | F | GG 1 |  |
| 67 | GSA-MA/WES + EPIC | Engel 1a | 2 | 0,5 | 6-10 | F | GG 1 | GG, PTPN11 |
| 68 | GSA-MA/WES | N/A | N/A | N/A | N/A | M | GG 1 |  |
| 69 | GSA-MA/WES | Engel 1a | 2 | 4 | 0-5 | F | GG 1 |  |
| 70 | GSA-MA/WES | Engel 1a | 2 | N/A | N/A | M | GG 1 |  |
| 71 | GSA-MA/WES | Engel 1a | 2 | 6 | 6-10 | F | GG 1 |  |
| 72 | GSA-MA/WES +<br>EPIC | Engel 1a | 2 | 0,25 | 6-10 | M | GG 1 | GG, PTPN11 |

